# Prediction of COVID-19 Active and Total Cases After a Fall and Rise of Cases

**DOI:** 10.1101/2020.07.02.20145045

**Authors:** Narayanan C. Viswanath

## Abstract

During the progress of the COVID-19, many countries have observed that their active cases tend to rise again after falling for some time. This may cause some mathematical models like the one discussed in [2] tend to make errors in the future prediction. We discuss a simple method to better the future prediction in such cases. This method is applied on the active and total cases data for the countries USA and Canada. In the case of Canada, the method succeeded in predicting the date when the active cases began to decrease. In the case of USA, a major improvement in prediction was observed when the method was applied: the predicted active and total cases are 1465602 and 2729015 for June 30; whereas the actual values are 1455400 and 2728856. We also give the active and total cases prediction for Canada and the USA for the first week of July 2020.

## Introduction

It can be observed that the path of active COVID-19 cases in the USA shows a decreasing tendency near May 30, 2020 (please refer [1]). A fit of the data between February 15 and May 20, 2020, based on a birth-and-death model (please refer [2]), suggests such a decreasing nature on May 28, 2020. This prediction remained the same, when we extended the data span as from February 15 to June 3, 2020. However in reality, the number of active cases began increasing from June 4, 2020 onwards. When we further extended the data span as first from February 15 to June 15, and then from February 15 to June 30, 2020 the above model predicted that a decrease in the active cases may occur from May 31 and June 13 respectively. These predictions turned out to be false as the number of active cases continued to increase from June 4 onwards. This suggested a poor fit of the data by the model, which predicted 2452094 total cases and 1207906 active cases on June 30 in place of actual 2728856 and 1455400 respectively, when fitted on data from February 15 to June 30, 2020.

A similar behavior was observed in the case of Canada, where a fit of the active cases data from February 15 to May 13, 2020 revealed that active cases may start decreasing from May 11, 2020. The actual data shows that there is a slight decrease in the active cases for three days starting from May 11 onwards. The active cases show an increasing nature from May 14 onwards and a continuous decrease occurs only from May 30 onwards. A fit of the data from February 15 to June 3, 2020 suggests a decrease in the active cases from May 17 onwards; whereas a fit of the data from February 15 to June 30, 2020 suggests this date to be May 20. This suggests that the fit is not very accurate: in place of actual 104204 total and 28019 active cases on June 30, the model (please refer [2]) predicted 105430 and 22341. However the inaccuracy in the fit is not that significant as in the case of the US. Canada’s cases being small in number when compared to the US may be the reason for this.

In the present study, we investigate how the model in [2] can be applied to fit a data where the active cases show a tendency to fall before rising further.

## Methods

The model is the same as the one defined by equations (8) and (9) in [2]. The method is to start the fitting from that point in time where the data begins to rise after falling. For example in the case of USA, we start the fitting from June 4, 2020. The active and total cases on this date are 1101255 and 1943347 respectively. For fitting the birth-and-death model, we take the active cases as given in [1], but starting from 1101255 (June 4 value). However, for equations (8) and (9) to be meaningful, we must take the total cases as starting from 1101255 itself. To achieve this, we subtract 1943347 and add 1101255 to the entire total cases data starting from June 4 onwards. While making predictions using the model fitted on the above data, we have to do the reverse procedure; that is we have to add 1943347 and subtract 1101255 to the predicted total cases.

## Results

### The United States of America

Figure 1 shows the comparison of the predicted and actual values by considering the entire data for the country USA from February 15 to June 30, 2020. It shows that the fitted model deviated much from the actual data. Figure 2 shows the comparison of the predicted and the actual values obtained by fitting the model as described in the methods section to the data from June 4 to June 30. The parameters that led to Figure 2 are given in Table 1. It should be noted that one must add 1943347 and subtract 1101255 to the total cases computed using the parameters in the table. Figure 2 clearly shows a better prediction for the above period. The predicted active and total cases by the new model for June 30 are 1465602 and 2729015, which are close to the actual values 1455400 and 2728856. This indicates the effectiveness of the new method discussed here. Table 2 contains the active and total cases in USA as predicted using the parameters given in Table 1 for the next 7 days; that is from July 1 to July 7.

**Figure 1.**
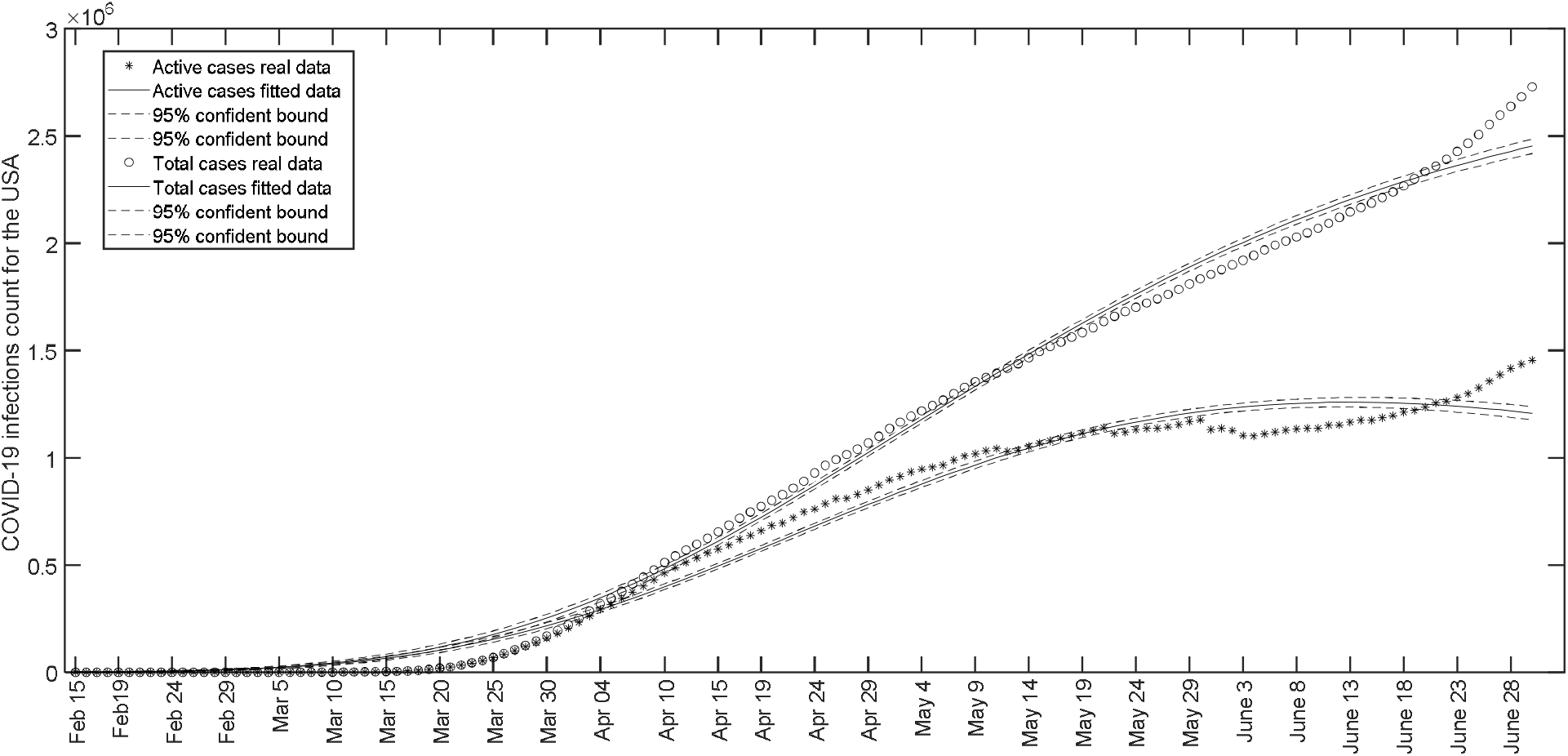
Comparison of the fitted values of active and total cases with the actual values for USA based on the data from February 15 to June 30, 2020.

**Figure 2.**
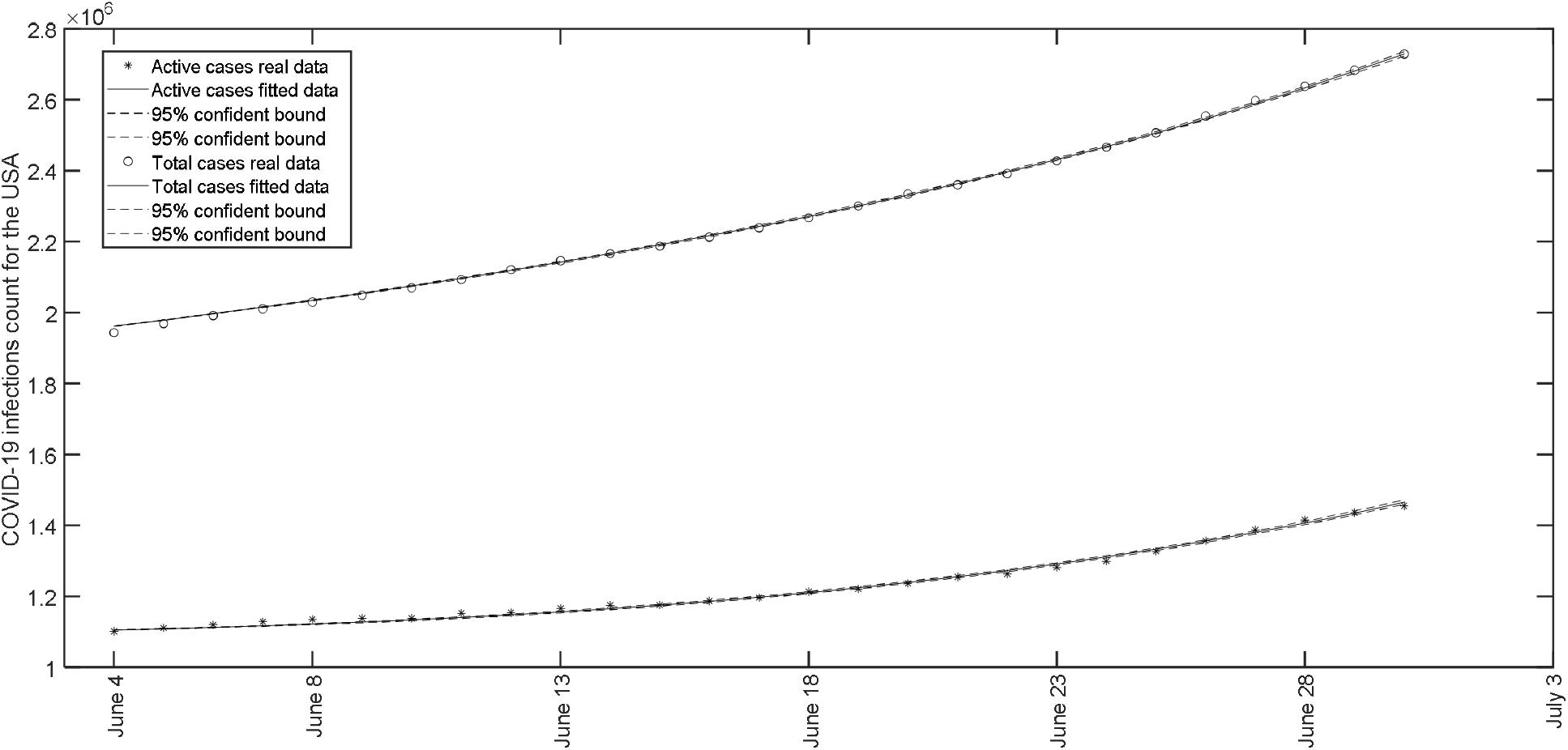
Comparison of the fitted values (when fitting was done as described in the methods section) of active and total cases with the actual values for USA based on the data from June 4 to June 30, 2020.

**Table 1.** Parameters for the fit of active and total cases for USA based on the data from June 4 to June 30, 2020.

| Parameter | a | b | $\mu$ | $N(0)$ |
| --- | --- | --- | --- | --- |
| Value | 0.014934868 | -0.030856395 | 0.012773050 | 1.102615554555564 |
| 95% Confidence Bounds | (0.0146, 0.015) | (-0.032, -0.029) | (0.0126, 0.01298) | [1.1, 1.1] |

**Table 2.** Prediction of the active and total cases for USA in the first week of July, 2020 using the parameters in Table 1.

| Date | Actives cases | Total cases |
| --- | --- | --- |
| July 1 | 1498382 | 2780722 |
| 2 | 1533572 | 2835273 |
| 3 | 1571359 | 2892887 |
| 4 | 1611951 | 2953806 |
| 5 | 1655576 | 3018296 |
| 6 | 1702486 | 3086649 |
| July 7 | 1752961 | 3159188 |

### Canada

In the case of Canada, Figure 3 shows that a fit (applying the model in [2]) of the actual and total cases during February 15 to June 30 is not as bad as in the case of USA. The future predictions are also good. However the fit predicts that actual cases may begin to decrease from May 20 onwards; whereas in reality this happens from May 30 only. Therefore, we apply the new method to check whether better prediction could be achieved. Since the active cases in Canada starts increasing from May 14, we consider a fit of the data from May 14 to June 30, 2020. The number of active and total cases on May 14 is 31838 and 73401 respectively. Therefore we have to subtract 73401 and add 31838 to the entire total cases data for Canada from May 14 to June 30 before fitting. The active cases data can be used without any change. Figure 4 shows the resulting fit of equations (8) and (9) in [2] to the active and total cases. Table 3 shows the parameters for the fit. Figure 4 shows a better fit of the data, compared to the Figure 3. The fitted model predicts the active and total cases for June 30 as 25969 and 104049 respectively; whereas the actual values are 28019 and 104204. Regarding the point where the active cases begin to decrease, the new model predicts it can happen from May 29, which is very close to the actual value May 30. This can be observed from Figure 4 also. In Table 4, we provide the prediction of active and total cases in Canada in the first week of July.

**Figure 3.**
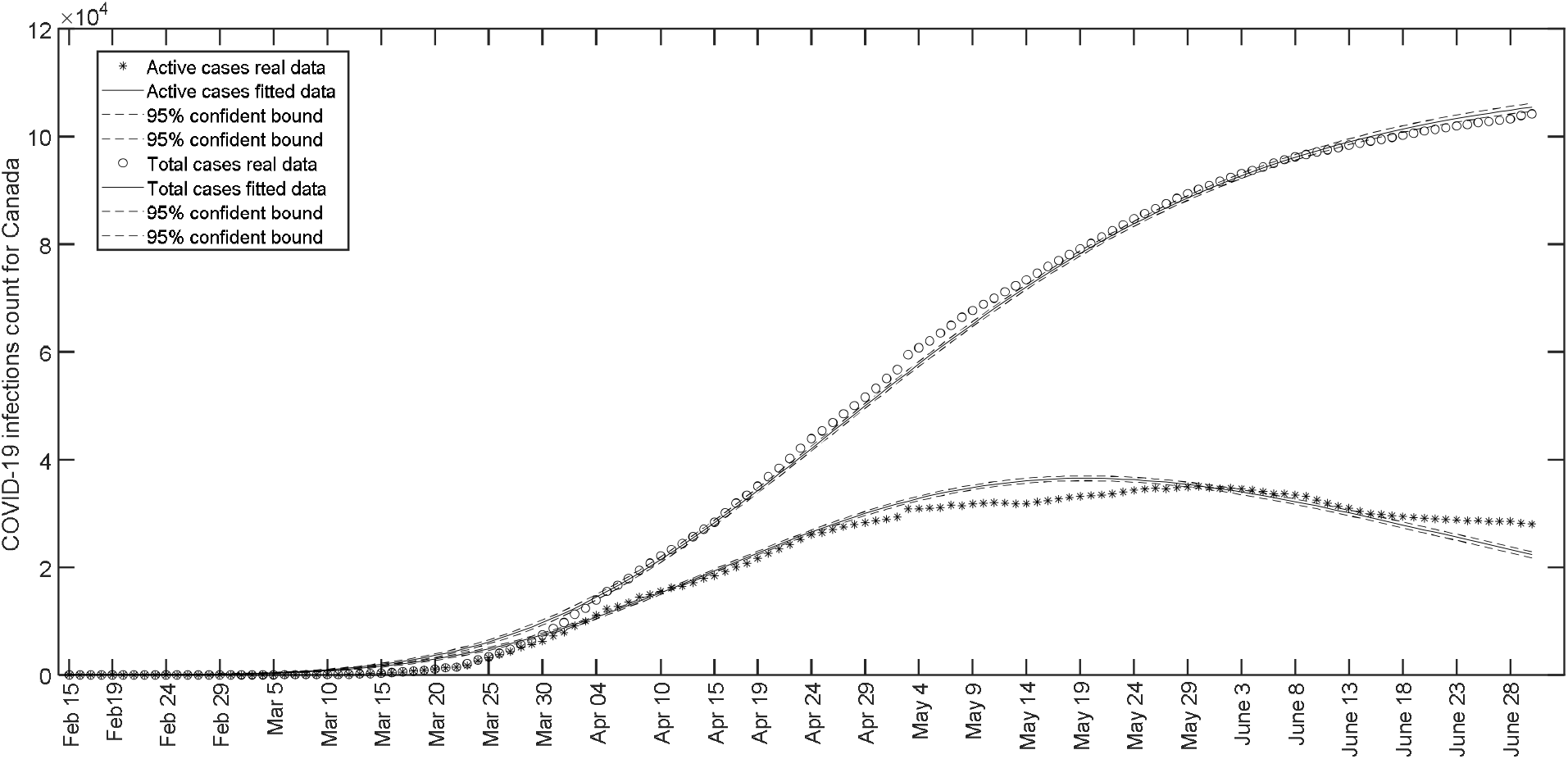
Comparison of the fitted values of active and total cases with the actual values for Canada based on the data from February 15 to June 30, 2020.

**Figure 4.**
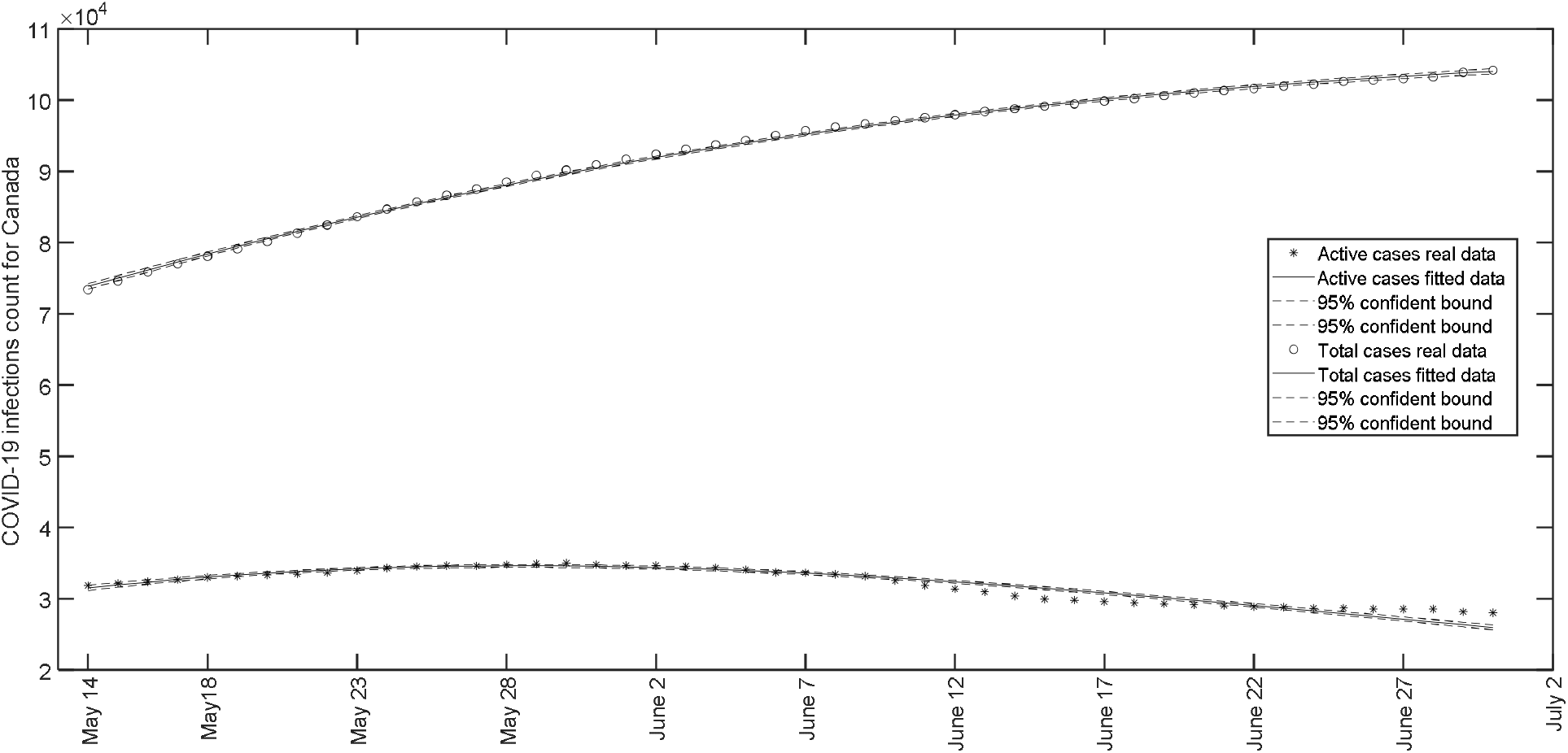
Comparison of the fitted values (when fitting was done as described in the methods section) of active and total cases with the actual values for Canada based on the data from May 14 to June 30, 2020.

**Table 3.**
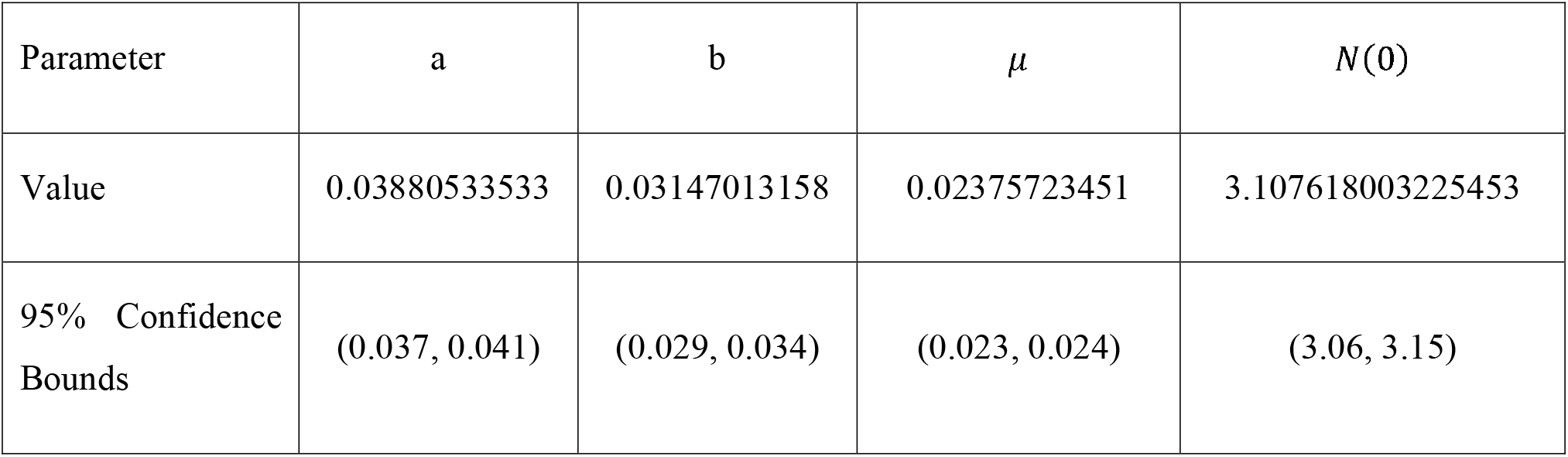
Parameters for the fit of active and total cases for Canada based on the data from May 14 to June 30, 2020.

| Parameter | a | b | $\mu$ | $N(0)$ |
| --- | --- | --- | --- | --- |
| Value | 0.03880533533 | 0.03147013158 | 0.02375723451 | 3.107618003225453 |
| 95% Confidence Bounds | (0.037, 0.041) | (0.029, 0.034) | (0.023, 0.024) | (3.06, 3.15) |

**Table 4.**
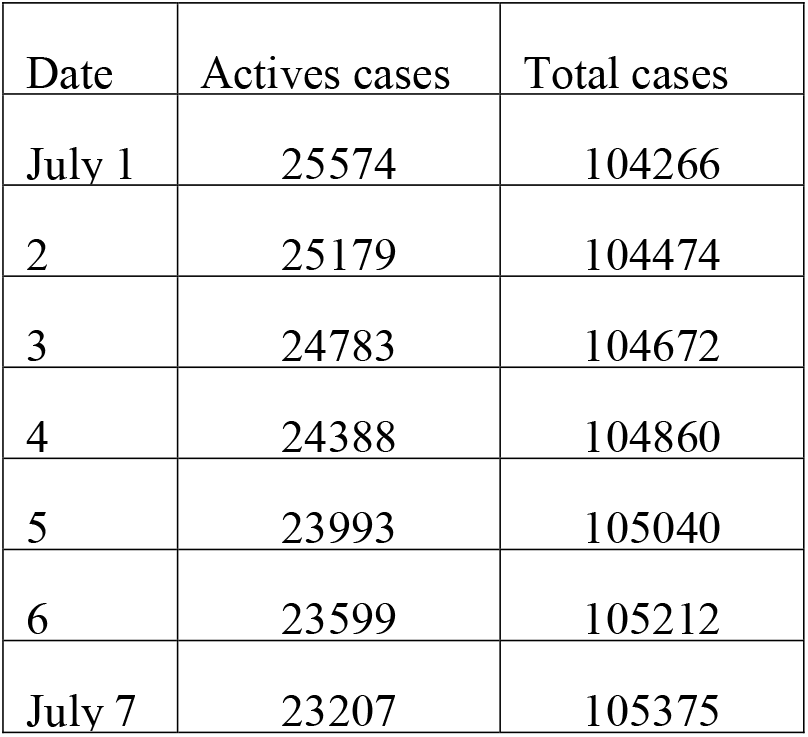
Prediction of the active and total cases for Canada in the first week of June, 2020 using the parameters in Table 3.

| Date | Actives cases | Total cases |
| --- | --- | --- |
| July 1 | 25574 | 104266 |
| 2 | 25179 | 104474 |
| 3 | 24783 | 104672 |
| 4 | 24388 | 104860 |
| 5 | 23993 | 105040 |
| 6 | 23599 | 105212 |
| July 7 | 23207 | 105375 |

## Data Availability

Data is available from the website
https://www.worldometers.info/

## Notes

### Competing Interest Statement

The authors have declared no competing interest.

### Funding Statement

No funding was received for the current study

### Author Declarations

Government Engineering College, Thrissur, Kerala, India

## References

1. Worldometers. Total Coronavirus Cases in the United States. https://www.worldometers.info/coronavirus/country/us. (June 30, 2020)

2. N C Viswanath. Analysis and Prediction of COVID-19 characteristics using a birth-and-death model. doi: https://doi.org/10.1101/2020.06.23.20138719 (2020)

